# Patient prioritisation methods to shorten waiting times for elective surgery: a systematic review of how to improve access to surgery

**DOI:** 10.1101/2021.02.18.21252033

**Authors:** Dimuthu Rathnayake, Mike Clarke, Viraj Jayasinghe

## Abstract

**Background:** Concern about long waiting times for elective surgeries is not a recent phenomenon, but it has been heightened by the impact of the COVID-19 pandemic and its associated measures. One way to alleviate the problem might be to use prioritisation methods for patients on the waiting list and a wide range of research is available on such methods. However, significant variations and inconsistencies have been reported in prioritisation protocols from various specialties, institutions, and health systems. To bridge the evidence gap in existing literature, this comprehensive systematic review will synthesise global evidence on policy strategies with a unique insight to patient prioritisation methods to reduce waiting times for elective surgeries. This will provide evidence that might help with the tremendous burden of surgical disease that is now apparent in many countries because of operations that were delayed or cancelled due to the COVID-19 pandemic and inform policy for sustainable healthcare management systems.

**Methods:** We searched PubMed, EMBASE, SCOPUS, Web of Science, and the Cochrane Library, with our most recent searches in January 2020. Articles published after 2013 on major elective surgery lists of adult patients were eligible, but cancer and cancer-related surgeries were excluded. Both randomised and non-randomised studies were eligible and the quality of studies was assessed with ROBINS-I and CASP tools. We registered the review in PROSPERO (CRD42019158455) and reported it in accordance with the PRISMA statement.

**Results:** The electronic search in five bibliographic databases yielded 7543 records (PubMed, EMBASE, SCOPUS, Web of Science, and Cochrane) and 17 eligible articles were identified in the screening. There were four quasi-experimental studies, 11 observational studies and two systematic reviews. These demonstrated moderate to low risk of bias in their research methods. Three studies tested generic approaches using common prioritisation systems for all elective surgeries in common. The other studies assessed specific prioritisation approaches for re-ordering the waiting list for a particular surgical specialty.

**Conclusions:** Explicit prioritisation tools with a standardised scoring system based on clear evidence-based criteria are likely to reduce waiting times and improve equitable access to health care. Multiple attributes need to be considered in defining a fair prioritisation system to overcome limitations with local variations and discriminations. Collating evidence from a diverse body of research provides a single framework to improve the quality and efficiency of elective surgical care provision in a variety of health settings. Universal prioritisation tools with vertical and horizontal equity would help with re-ordering patients on waiting lists for elective surgery and reduce waiting times.

## INTRODUCTION

Concern about long waiting times for elective surgeries is not a recent phenomenon, but it has been heightened by the impact of the COVID-19 pandemic and its associated measures. Waiting times are a key performance indicator for many healthcare systems and waiting-time targets are used to encourage improved performance in healthcare institutions to deliver high-quality care without unnecessary delay (1). The COVID-19 pandemic has already added considerably to this challenge, leading to a considerable increase in the pressure on healthcare institutions to clear patients in clinics. The healthcare system slowdown caused by the pandemic and its associated measures has had a substantial impact on elective services (2) and efficient allocation of elective surgical resources are more critical than ever in the business as unusual (BAUU) future for healthcare systems and institutions (3). There is also a need for strategies implemented during the pandemic to be continually integrated into hospital practices, to minimise the risk of transmission of the coronavirus (4).

Waiting lists are considered as a non-price rationing mechanism for coping with excess demand. Long waiting times are associated with many adverse effects, including a higher risk of death and serious complications for patients, especially adults (5-10). Consequently, waiting times for elective surgeries are a major policy concern in many countries, especially for health systems operated with public funds (11). Despite increased funding in recent years, the demand for many elective surgeries exerts a substantial challenge, which was growing even before the COVID-19 pandemic (12). The negative impact of patient waiting time on cost-effectiveness in economic evaluations has also found to be non-reversible (13-15) and there is a need for economic evaluations, adapted to outbreak situations (16), to estimate the specific impact of COVID-19 pandemic on the pre-existing crisis of longer waiting times.

A large amount of literature is available on a large array of methods to reduce patient waiting times for elective surgeries. The research focus has shifted recently to individual strategies, rather than system-wide approaches. One of these strategies is to prioritise patients, such that the waiting list is re-ordered. Although prioritisation does not always reduce total waiting time for all patients, it allows those patients in the greatest need to be treated first.

Prioritisation processes are used in many countries to order the queue of patients for high-demand surgeries, and many research studies have demonstrated promising results for various patient prioritisation methods, but there are few systematic reviews of the effects of these methods. As an example, a systematic review done in 2003 on patient prioritisation for elective surgery sought to determine the basis of ethical approaches used in different prioritisation tools (17), and other systematic reviews have analysed different approaches to reduce waiting times for elective surgery (18-20). To bridge the evidence gap in the current literature, this systematic review seeks to identify opportunities to shorten waiting times for elective surgery with a particular focus on patient prioritisation methods. It seeks to answer the following “W*hat are the effective patient prioritisation methods to reduce waiting times for elective surgeries and how consistent were those results to different elective surgical specialties, institutions, and health systems?”*

## METHODS

This review is one of the sub-reviews in a major systematic review conducted with a broader search to support a holistic approach to finding solutions for long waiting times for elective surgery. It assesses patient prioritisation as one strategy among a much wider scope of approaches. The full, portfolio review was registered in PROSPERO (CRD42019158455) and its PRISMA flow diagram is attached (Supplementary online file 1). The broad scope allows for the inclusion of all methods, strategies, and policies to reduce waiting times for elective surgery. For this sub-review, studies on patient prioritisation methods were deemed eligible, and it was conducted and reported according to the PRISMA statement and checklist (Supplementary online file 2). Given that healthcare system interventions are often tested in quasi-experimental studies or observational studies, rather than experimental studies (such as randomised trials), a range of study designs were eligible for this review. With this in mind and because the validity of the results of any systematic review of health system interventions is dependent on the use of relevant evidence and synthesis methods (21), we used design-specific tools to evaluate the risks of bias associated (22), as discussed below.

### Data sources for the portfolio review

We searched PubMed, EMBASE, SCOPUS, Web of Science, and the Cochrane Library using combinations of search terms. After pilot searches, we finalised a detailed search strategy which consisted of three sets of search terms without language restrictions. The searches were run from 14 December 2019 to 7 January 2020, for articles published from 1 January 2014 to December 2019. The search strategy used for PubMed is presented in Supplementary online file 3.

### Criteria for considering studies for the review

We included studies that investigated interventions and strategies intended to reduce waiting time for elective surgery. We included original research published in journal papers, reports, editorials, and literature reviews from the health sector, government, and related sectors. We included experimental, quasi-experimental, and observational studies, as well as systematic reviews published during 2014-2019. Qualitative and quantitative data were considered for data synthesis; but simulation and modelling studies were excluded, because they might not be a reliable guide to the effects in real-world scenarios.

Eligible participants are adult patients (≥18 years) registered for elective surgery. Patients undergoing emergency surgery and surgery for paediatric conditions were excluded. Studies of waiting times for day surgery or ambulatory surgery were eligible. Studies with patients waiting for all types of major elective surgery were eligible, except for those of patients waiting for cancer or cancer-related surgeries. The surgeries require penetration of a body cavity are considered as major surgeries and all surgeries of abdomen, chest or cranium are considered major surgeries. Minor surgeries are generally superficial and do not require penetration of a body cavities (23). Although most eye surgeries are minor surgeries, studies on eye surgery lists were included as an exception, because these have some of the longest surgery waiting lists in many countries (24). For this review, where a study was based in a clinic or outpatient department, we required that the investigation was targeted on the prioritisation of patients registered for elective surgery, rather than patients waiting for other procedures.

### Article selection and data extraction

DR and VJ to select relevant articles checked the title and the abstract of retrieved citations. Articles that were deemed potentially eligible based on their title or abstract were retrieved in full and assessed for eligibility and relevance. Each potentially eligible article was discussed with the third reviewer (MC) and agreement was reached on inclusion or exclusion.

### Synthesis of results

The portfolio review includes a wider scope and data synthesis for this review was carried out as a single subgroup analysis of that wide series of systematic reviews. Meta-analyses were not applicable for this review because of the heterogeneity in study designs and variability of the approaches to how the outcome of interest was measured. Instead, we planned a meta-synthesis with narrative analysis. Given the types of study that we identified, we used the ROBINS-I tool (25) for quality evaluation in non-randomized intervention studies and the CASP tool (Critical Appraisal of Skill Programme) for observational studies (26). We would have used the Cochrane Risk of Bias tool for randomized trials, but none were identified for this review. We used common criteria to report the overall quality of evidence in observational studies.

## RESULTS

The article screening process is shown as a PRISMA flowchart (Figure 1). The electronic search for the full review yielded 7543 records from the five bibliographic databases. This reduced to 5346 after deduplication in EndNote citation management software. During the title and abstract screening process, 362 potentially relevant citations were selected, and this was reduced to 196 articles after full article screening for the extended scope of the full, portfolio review. Of these, 105 simulation and modelling studies were rejected at this stage. After grouping the citations to different strategies for the same intended outcome of reducing waiting times, 17 articles were judged eligible for this sub-review because their major emphasis was on methods for prioritising patients to reduce waiting time for elective surgery.

**Figure 1.**
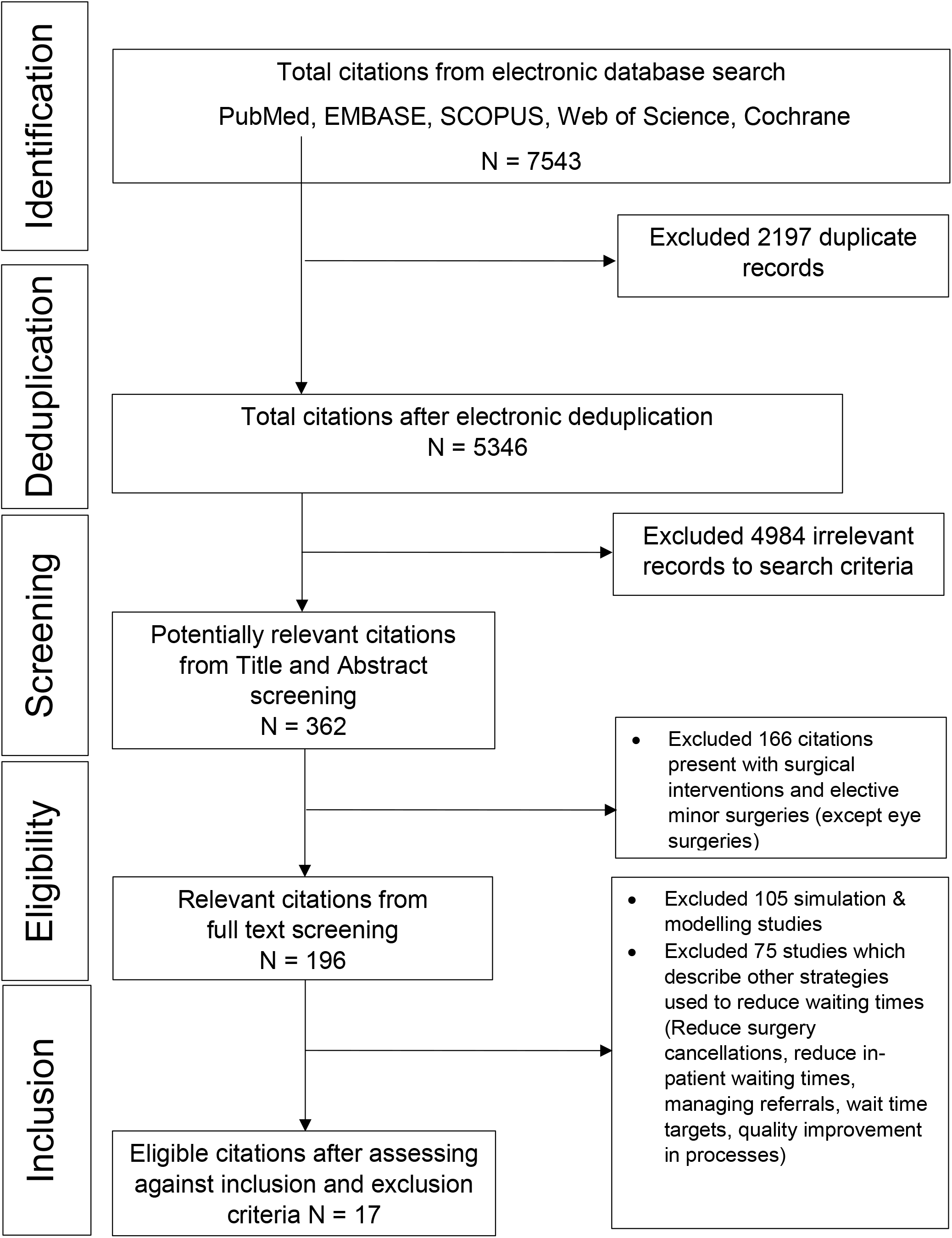
PRISMA flow diagram for eligible article selection for the systematic review

Our primary outcome variable is waiting time, which was defined as the period between a surgeon placing a patient on the waiting list for a particular elective surgery and the day that the surgery is performed. In total, we included 17 studies that were published between 2014 and 2019: four quasi-experimental studies, 11 observational studies and two systematic reviews. A summary is presented in Table 1. Of the 15 original studies (excluding the two systematic reviews), ten described prioritisation as a health system-wide approach, and five tested patient prioritisation as an institutional measure, investigating the association of waiting time with an explicit prioritisation guideline.

**Table 1.**
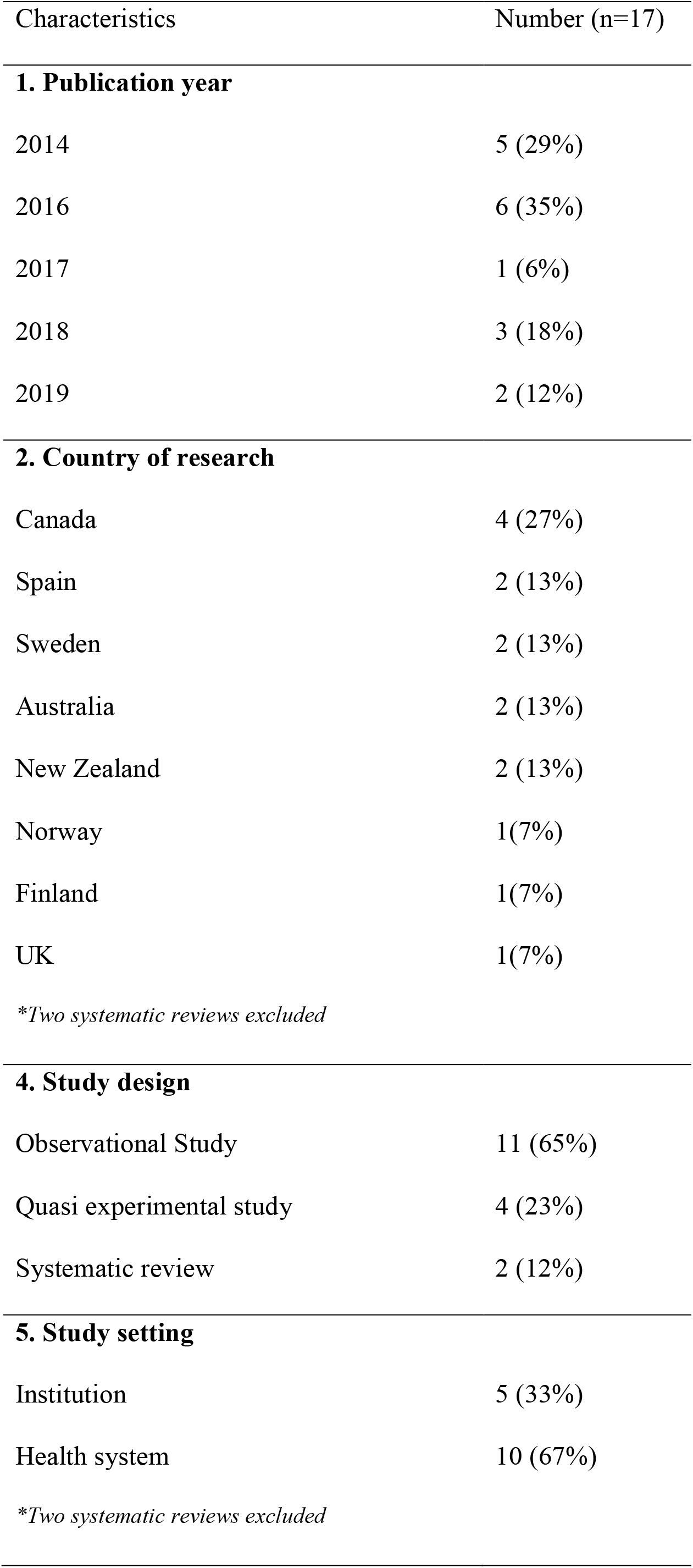
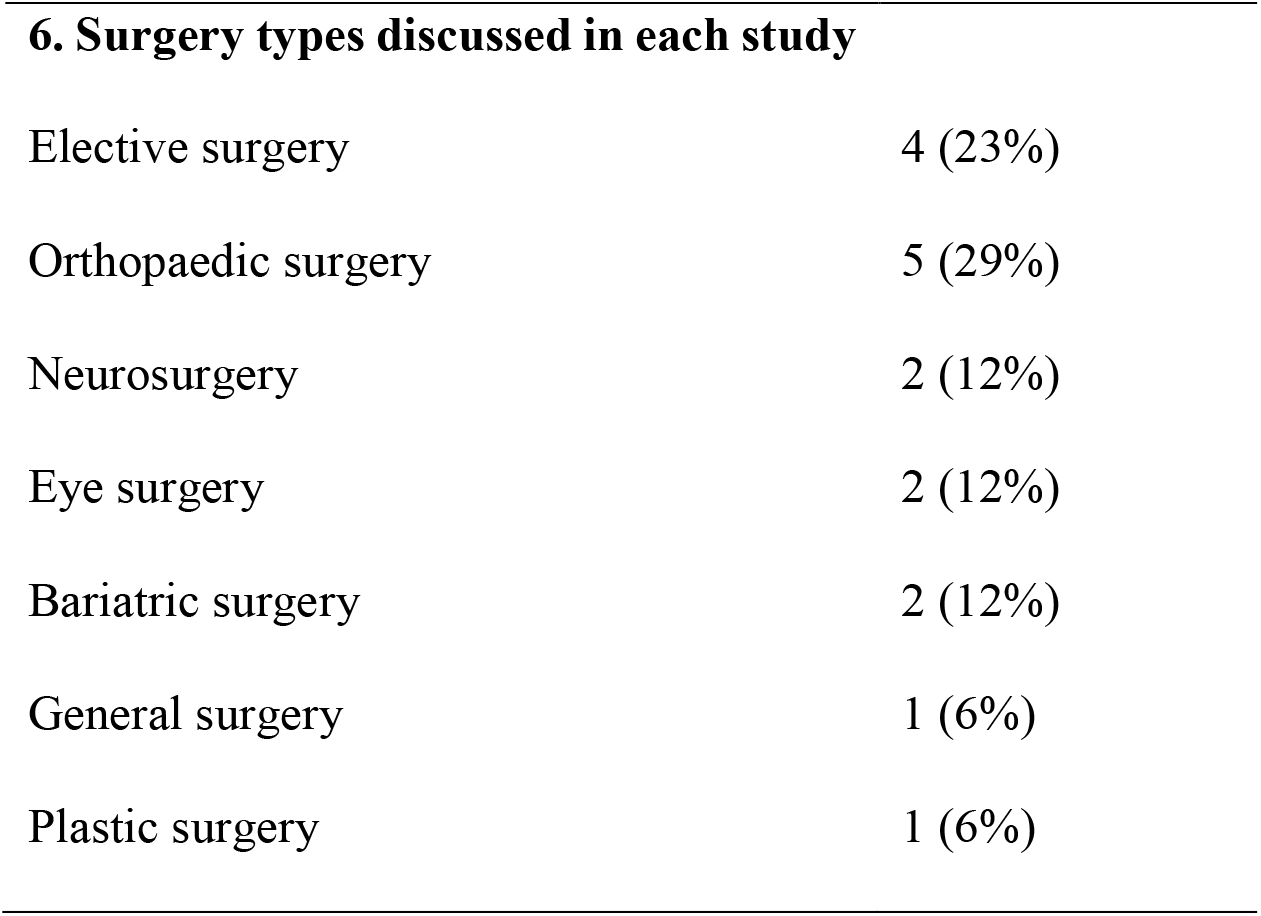
Summary of characteristics in included studies

| Characteristics | Number (n=17) |
| --- | --- |
| **1. Publication year** |  |
| 2014 | 5 (29%) |
| 2016 | 6 (35%) |
| 2017 | 1 (6%) |
| 2018 | 3 (18%) |
| 2019 | 2 (12%) |
| **2. Country of research** |  |
| Canada | 4 (27%) |
| Spain | 2 (13%) |
| Sweden | 2 (13%) |
| Australia | 2 (13%) |
| New Zealand | 2 (13%) |
| Norway | 1(7%) |
| Finland | 1(7%) |
| UK | 1(7%) |
| *\*Two systematic reviews excluded* |  |
| **4. Study design** |  |
| Observational Study | 11 (65%) |
| Quasi experimental study | 4 (23%) |
| Systematic review | 2 (12%) |
| **5. Study setting** |  |
| Institution | 5 (33%) |
| Health system | 10 (67%) |
| *\*Two systematic reviews excluded* |  |
| **6. Surgery types discussed in each study** |  |
| Elective surgery | 4 (23%) |
| Orthopaedic surgery | 5 (29%) |
| Neurosurgery | 2 (12%) |
| Eye surgery | 2 (12%) |
| Bariatric surgery | 2 (12%) |
| General surgery | 1 (6%) |
| Plastic surgery | 1 (6%) |

### Summary of included studies

The characteristics of the 17 included studies are summarised below and further details are given in Table 2.

**Table 2.**
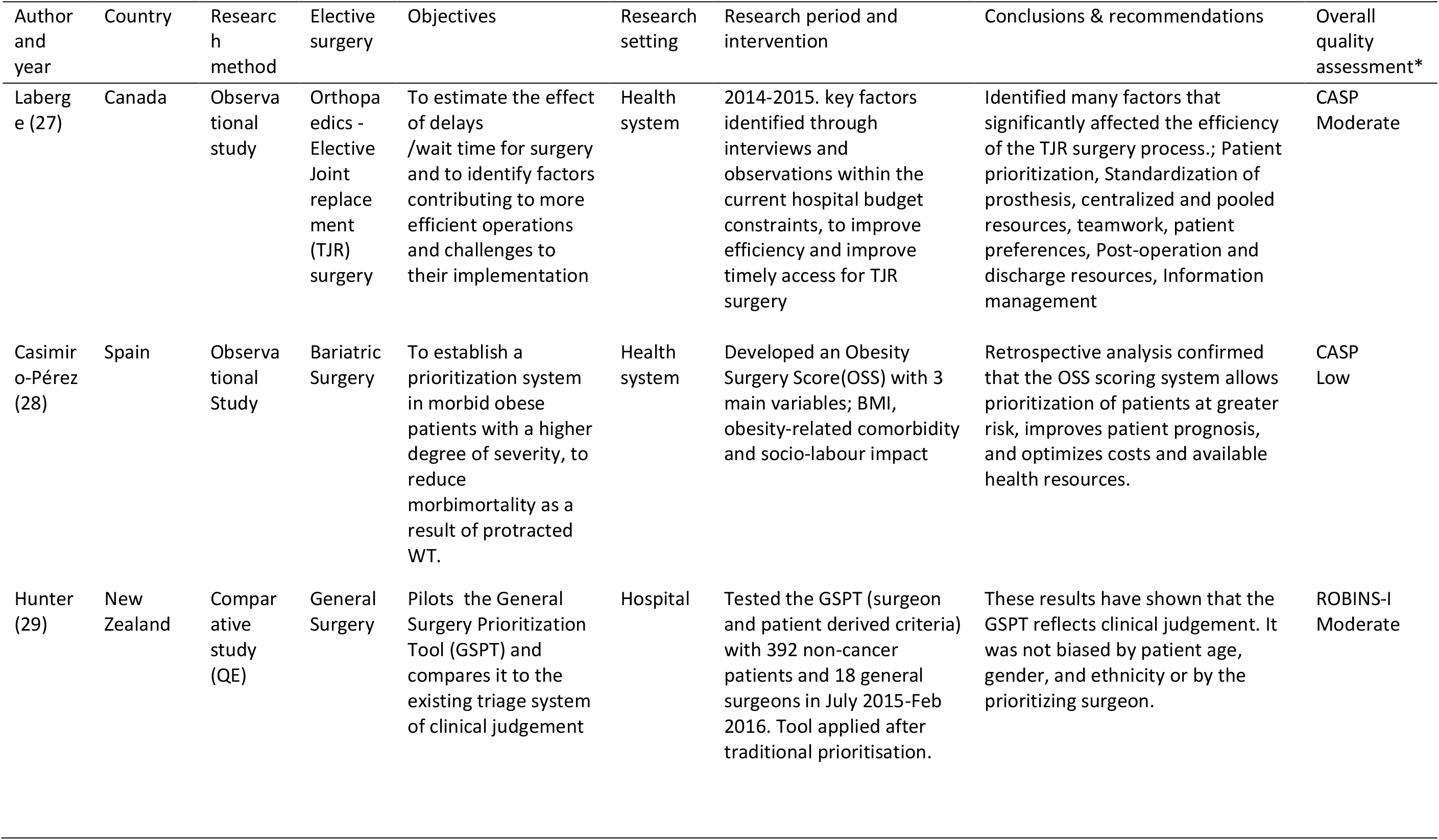
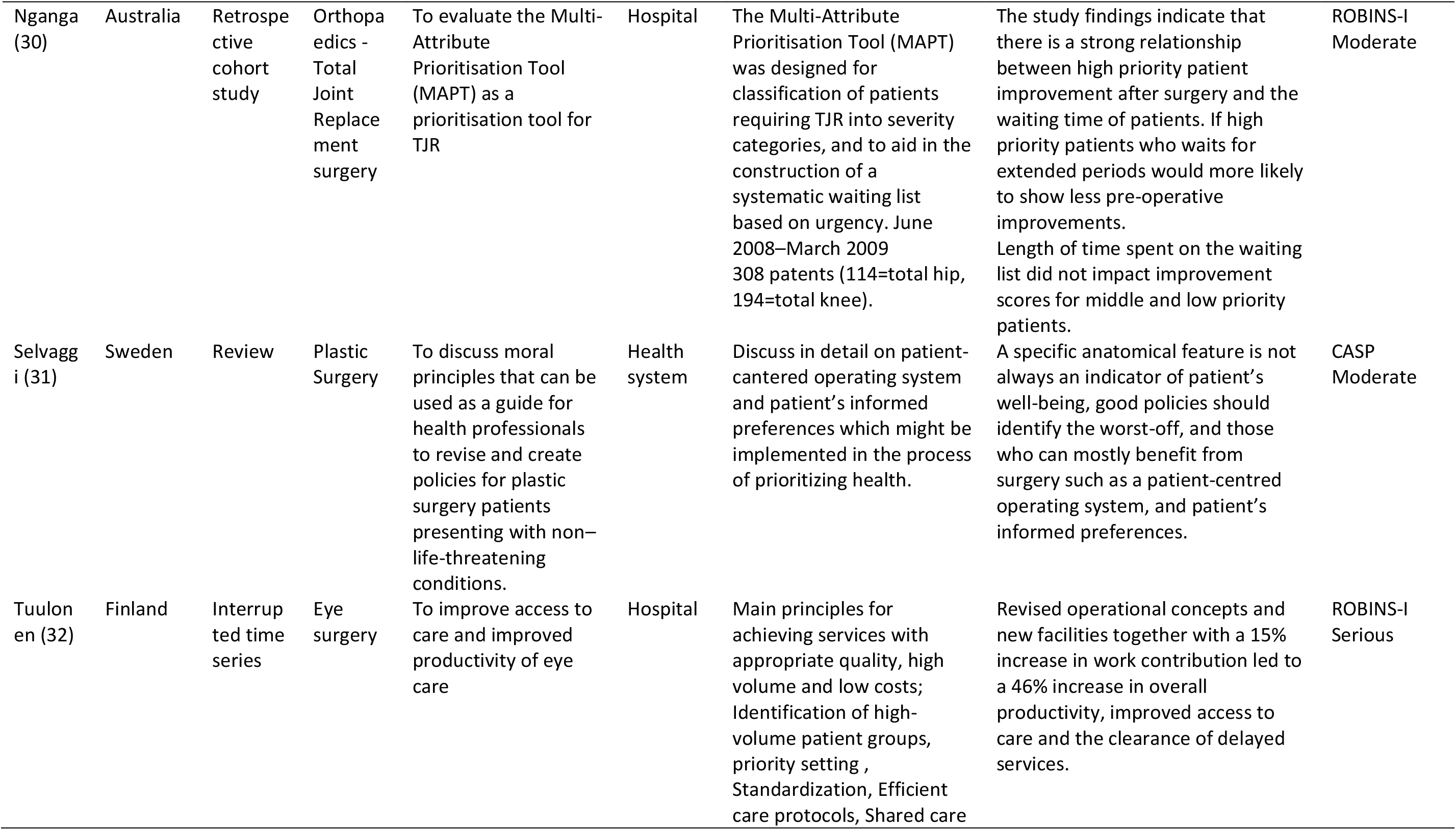
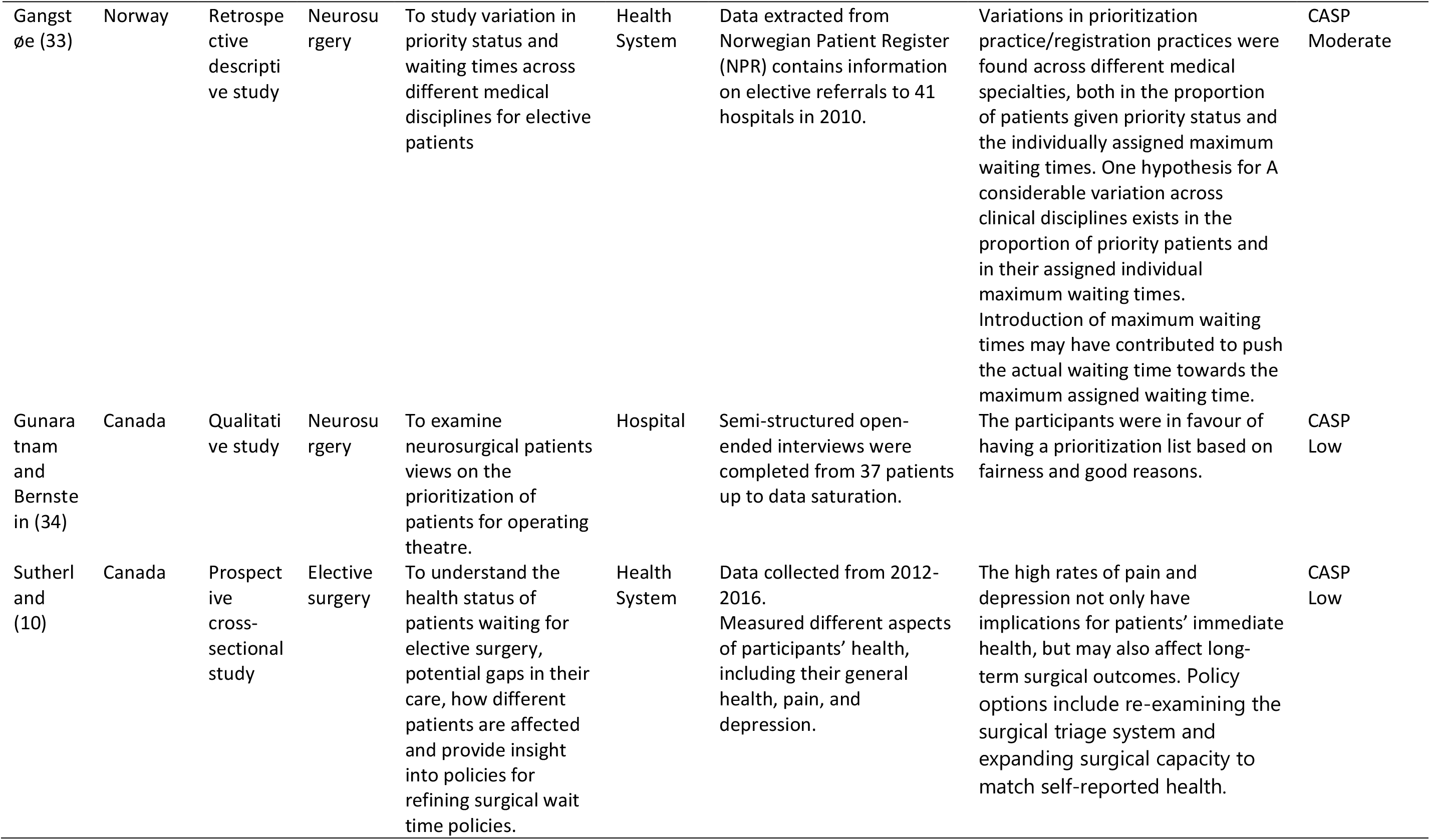
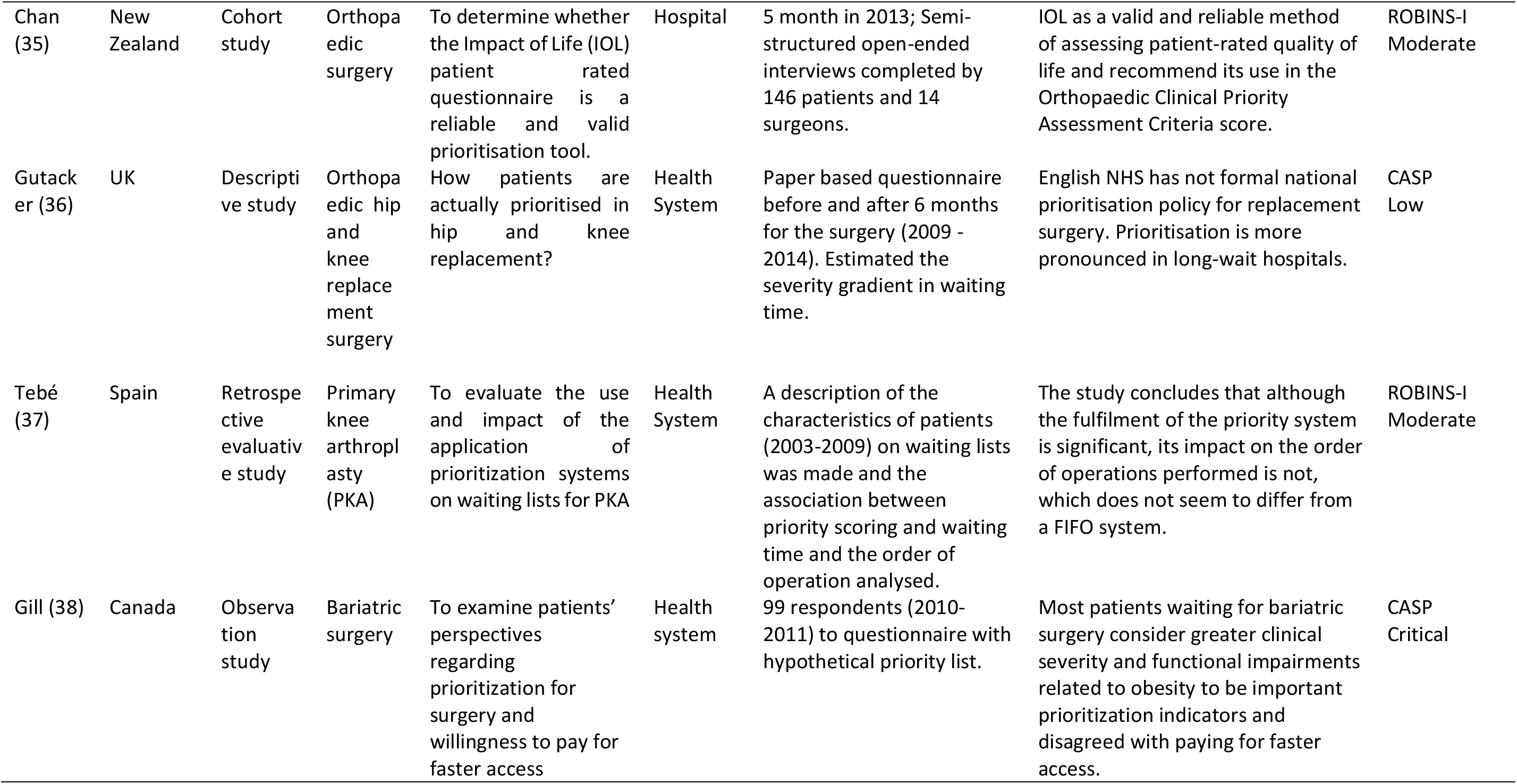
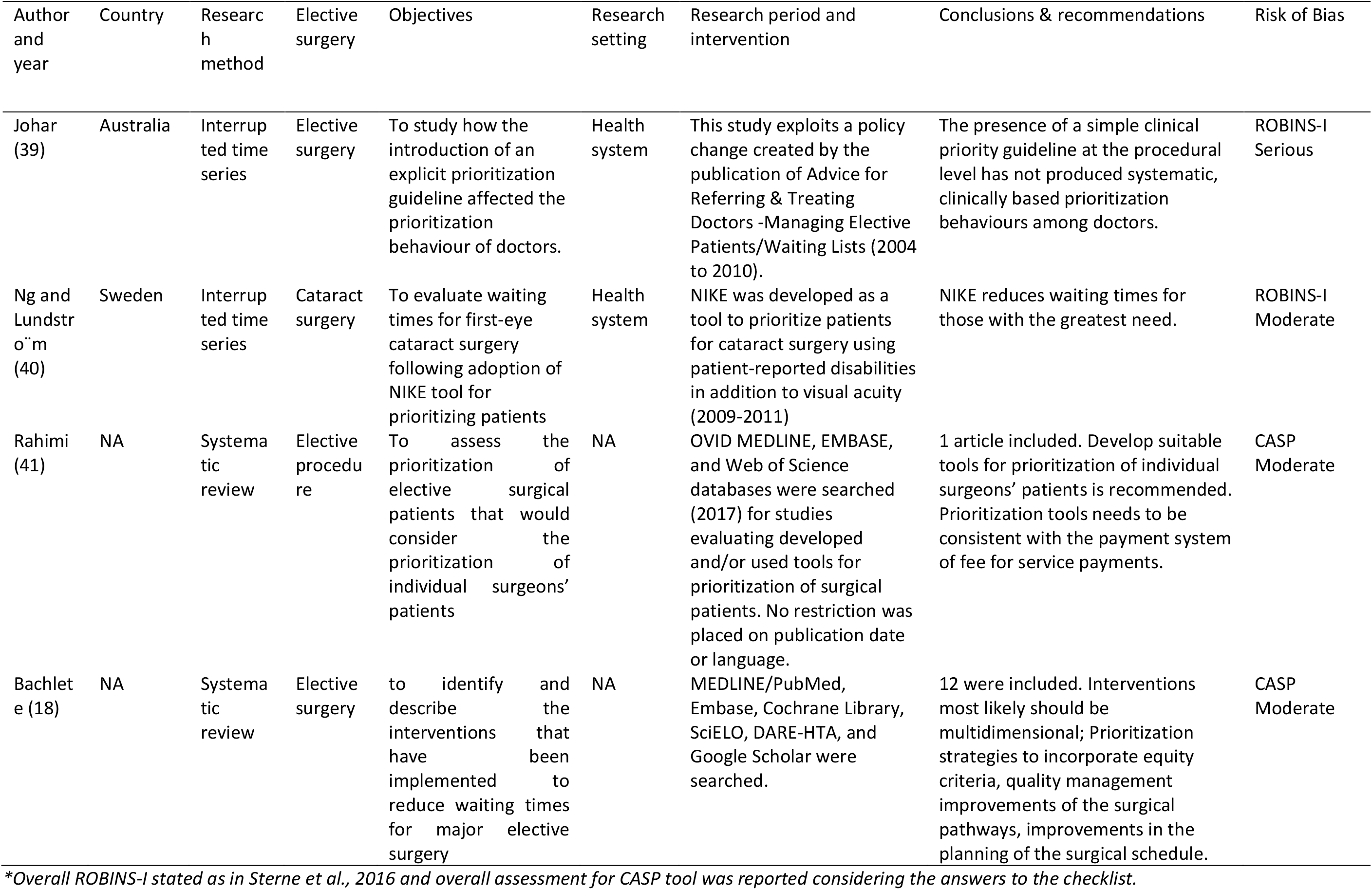
Descriptive characteristics of the seventeen studies included for the review and their overall quality assessment

Laberge (27) investigated the effects of delays and waiting times for total joint replacement (TJR) surgery using observational study methods. The study was conducted in a university-affiliated hospital in 2014-2015 in Canada. The results identified multiple factors that significantly affected the efficiency of the TJR surgery process, with proper patient prioritisation being among the recommendations to investigate, along with process engineering with health services and management research.

Casimiro-Perez (28) reported a clinical prioritisation system for patients on a waiting list for bariatric surgery, called the Obesity Surgery Score (OSS), in Spain. This observational study was conducted with the OSS for different hospital waiting lists and it concluded that applying a structured prioritisation system such as OSS to bariatric surgery waiting lists might improve patient prognosis, timeliness of surgery, and cost optimization.

Hunter (29) reported a quasi-experimental study to pilot a General Surgery Prioritisation Tool (GSPT) with an existing triage system of clinical judgment, in New Zealand. A cohort of 392 patients added to a waiting list between July 2015 and February 2016 was selected for the intervention. The tool was found to be clinically reliable, with the advantage of being a more explicit process.

Nganga (30) reported a cohort study from Australia, including 308 patients undergoing total hip or knee arthroplasty from June 2008 to March 2009. The study examined pre-operative and post-operative Multi-Attribute Prioritisation Tool (MAPT) scores for these patients. The results indicated that the patients who were prioritised by the MAPT questionnaire had an improved MAPT score of zero or close to zero postoperatively.

Selvaggi (31) discussed moral principles that can be incorporated in prioritisation criteria and used to guide decision making by health professionals. It recommended the implementation of patient-informed preferences for plastic surgery patients presenting with non–life-threatening conditions in Sweden.

Tuulonen (32) reported an intervention study to improve access and productivity for eye care in Finland. Along with other strategies, patient prioritisation on the basis of patients’ risk for permanent visual disability increased the number of surgical procedures throughout the period (2014 to 2015).

Gangstoe (33) reported a cross-sectional study of the variation in priority status and waiting times across different medical disciplines for elective patients admitted to specialized services in Norway, in 2010. Considerable variation was found across medical specialties, with causes for variation often interpreted as differences in clinical judgment and capacity.

Gunaratnam (34) conducted a qualitative study to examine neurosurgical patients’ views on the prioritisation of patients for operating theatre in Canada in 2015. They interviewed 37 patients and concluded in favour of having a prioritisation list based on fair methodology.

Sutherland (10) conducted a cross-sectional study to report the health status of patients waiting for elective surgery and to provide insight into policies for refining surgical wait time policies. A total of 3759 patients responded to the survey questions. The findings indicated the need for non-surgical interventions while patients were waiting for elective surgery, pointing to a range of policy options for triaging elective surgery patients.

Chan (35) reported a cohort study to test the reliability and validity of the Impact on Life (IoL) patient-rated questionnaire for use in prioritising orthopaedic surgeries. 324 patients responded to the questionnaire over a 5-month period in 2013. The IoL was judged to be a relevant, comprehensive and user-friendly tool for use in the prioritisation of elective surgical services.

Gutacker (36) reported a descriptive study of how patients are prioritised in hip and knee replacement surgeries in the UK, during 2009 to 2014. The authors concluded that the factor of not practicing formal and explicit prioritisation system for elective surgeries in English NHS hospitals was one major cause for lengthening waiting lists for elective surgery.

Tebé (37) reported an evaluation of the impact of using a priority system for primary knee arthroplasty (PKA), in Spain. Data from a management registry in the period of 2003 to 2009 were analysed, with 67,403 patients included. The study concluded that although a prioritisation system for PKA had been implemented, it had no effect on the prioritisation of patients based on their severity.

Gill (38) reported an observational study that examined patients’ perspectives regarding prioritisation for bariatric surgery and the willingness to pay for faster access, in Canada. 99 patients responded (2010 to 2011) and most considered greater clinical severity and functional impairments related to obesity to be important prioritisation indicators and disagreed with paying for faster access.

Johar (39) reported an intervention study to investigate how the introduction of an explicit prioritisation guideline affected the prioritisation behaviour of doctors in Australia during 2004 to 2010. The study revealed that the presence of a simple clinical priority guideline at the procedural level did not produce systematic, clinically based prioritisation behaviours among doctors.

Ng and Lundstrom (40) reported waiting times for cataract surgery in Sweden following the adoption of the National Indications model for Cataract Extraction (NIKE) tool for prioritizing patients. There were 141,070 surgeries during the study period (2009 to 2011) and mean waiting times decreased across all NIKE groups, with an annual increase of surgery rate around 6%.

Rahimi (41) is a systematic review of studies for the prioritisation of elective surgical patients considering the prioritisation of individual surgeons’ patients. They searched OVID MEDLINE, EMBASE, and Web of Science in 2017. A single study was eligible form the screening and it was from the main author. This illustrated the use of prioritisation of patients among the waiting lists of two individual orthopaedic surgeons in a large teaching hospital in Iran.

Bachelet (18) is a scoping review of studies of interventions that have been implemented to reduce waiting times for major elective surgeries. They searched six electronic databases up to December 2017 and included 12 eligible studies. They assessed the quality of the evidence with EPOC (The Cochrane Effective Practice and Organisation of Care) and GRADE (Grades of Recommendation, Assessment, Development, and Evaluation) tools, and rated the overall quality of all studies as low. They concluded that there is a need for multidimensional interventions based on prioritisation strategies, and quality management improvements of the surgical pathways and improvements in the planning of the surgical schedule.

### Risk of bias in included studies

The quality of the individual studies that we included in this review was measured using two tools. In addition to the four intervention studies, we evaluated three of the other studies with ROBINS-I considering the longitudinal assessment of their outcome variable (29, 30, 35). The relevant CASP reading tool was used to evaluate the other ten studies.

#### ROBINS-I

We judged five studies to have an overall moderate quality ((29, 30, 35, 37, 40) and two studies to have a serious risk of bias in their methodology (32, 39). Details of ROBINS-I evaluation for each domain is shown in Figure 2 and Figure 3.

**Figure 2.**
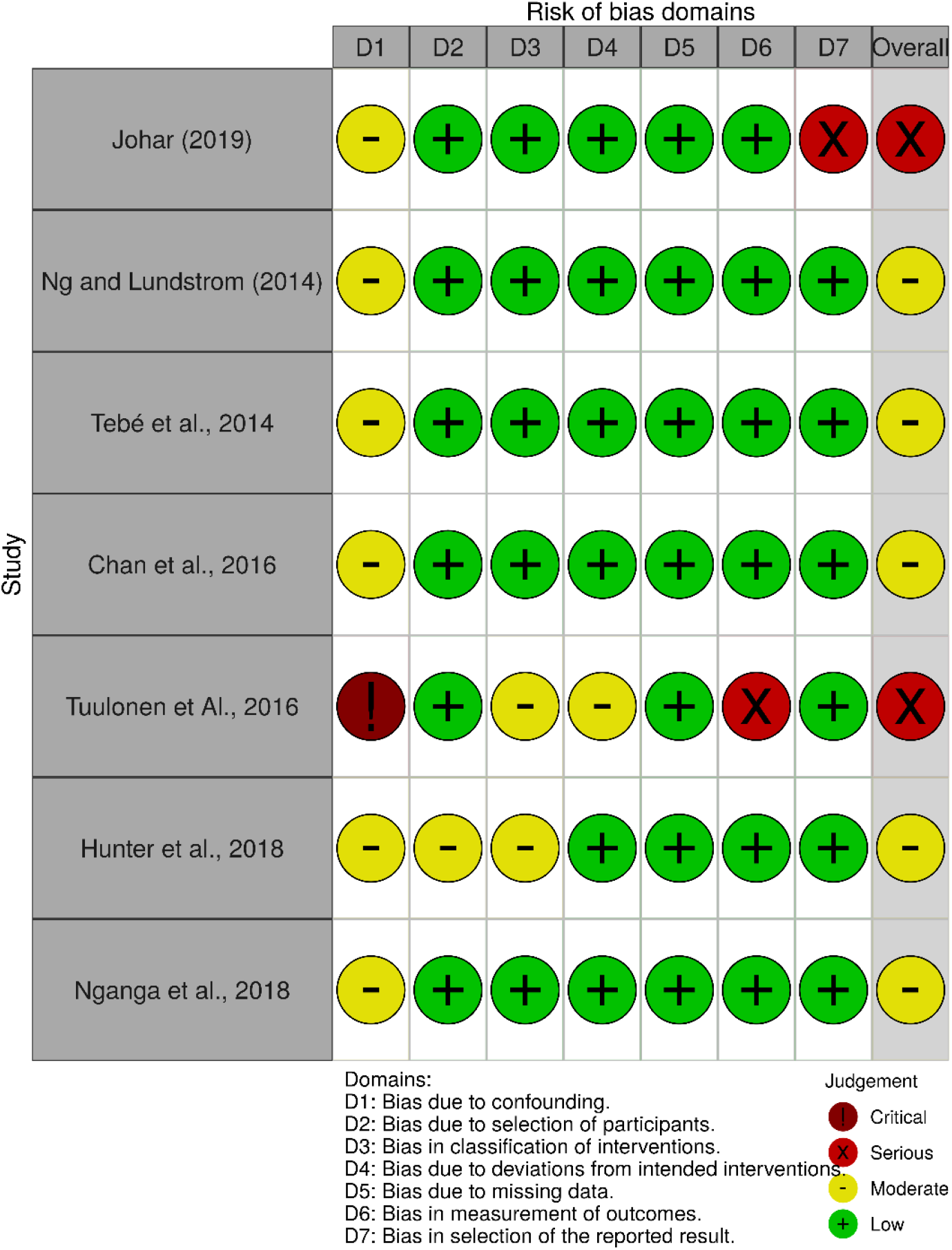
Traffic-light plot for the risk of bias domains in ROBINS-I for the selected studies.

**Figure 3.**
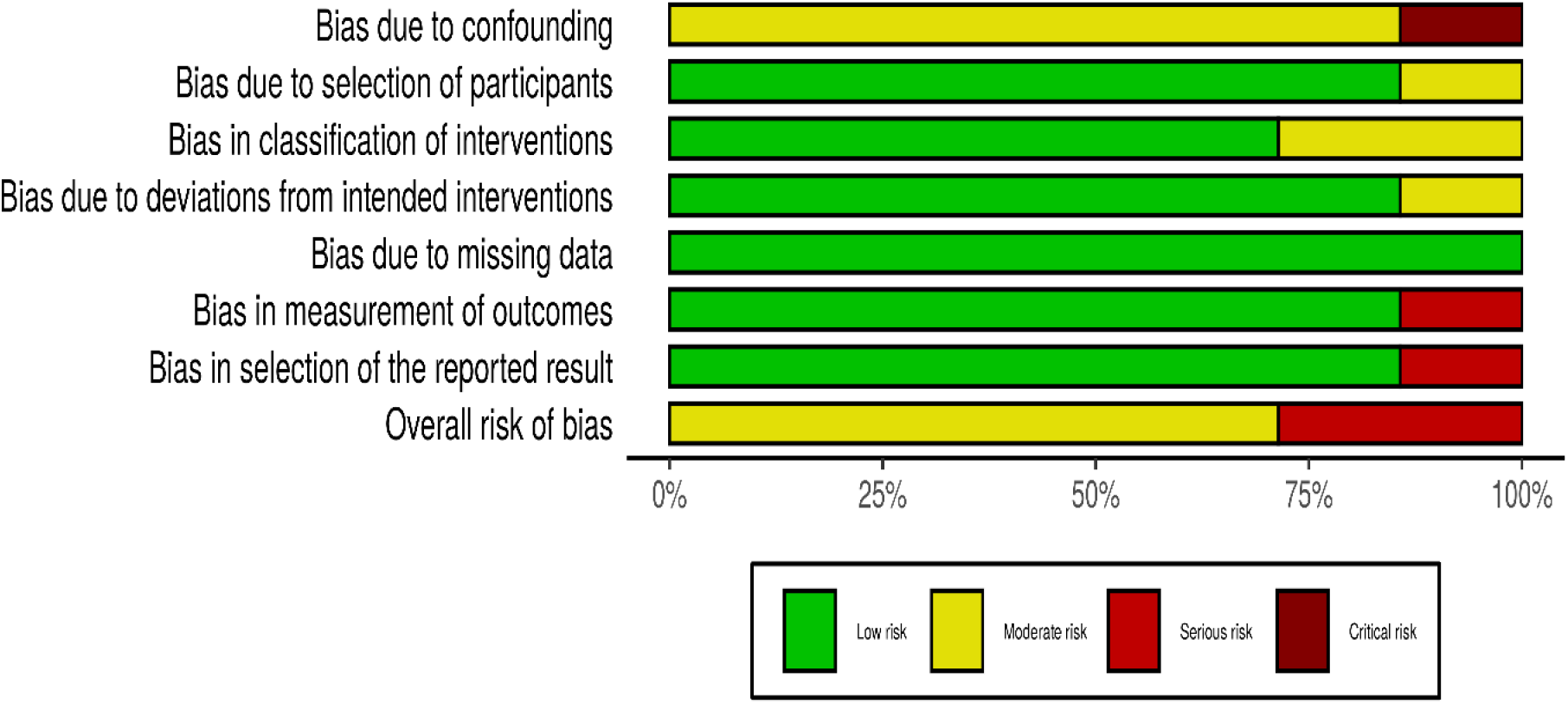
Bar diagram for each risk of bias domains in ROBINS-I for the selected studies.

All seven of these studies had inclusion and exclusion criteria for study participants, but none had used randomisation to allocate participants to the assessed prioritisation processes. We identified moderate risk of bias due to confounding for baseline characteristics of the comparison groups in all studies. One study had a critical risk of bias due to confounding because of the changing of the research setting during the intervention (moved to a new hospital building), although it reported change in pre and post stages of intervention measures (32). There was a low risk of bias in the selection of the participants in all studies because patients were recruited as entire blocks to each study. The bias in the classification of the intervention was low in all studies because the interventions were implemented in pre-post design methods, with participants unaware of the prioritisation scoring at the clinics. The introduced interventions are explicit prioritisation guidelines and most indicators are dependent on objective parameters. Therefore, there were no significant deviations reported from the intended intervention. There were no missing data reported in any of the studies, because waiting times were extracted from regular patient records in administrative documents.

However, one study had a serious risk of selection bias of reported results because of the limitation of using admission data only from those patients who had completed the waiting period. In this study, the number of patients who had not attended the admission was not reported (39). Outcome variables included in many studies were as intended, namely a measurement of the time spent on the waiting list, which was unbiased.

#### CASP tools

The main outcome variable of interest for this review is a time-to-event measurement, which required the follow up of a cohort of patients and various time measurements in these study participants. We applied the CASP checklist for cohort studies to six included studies (10, 27, 28, 33, 36, 38) and the CASP checklist for Qualitative studies to two studies (31, 34). The two systematic reviews were assessed using the CASP checklist for systematic reviews.

We assessed most studies to be of moderate to low risk of bias. The evidence from most studies is sufficient to support their results (10, 27, 31, 33, 34, 36, 41, 42). However, one study demonstrated poor study methodology because confounding factors were not considered when recruiting study participants or in the data analysis (43).

## DISCUSSION

Seventeen research studies were eligible for this narrative synthesis. Meta-analyses were not possible or appropriate because of the substantial heterogeneity across the studies. Prioritisation strategies for waiting lists incorporate equity criteria (44) and, so, as a first step in our narrative synthesis, we focused on the levels of implementation of the prioritisation systems in each study. Many prioritisation scores were recommended to be used for individual disciplines and there were two apparent approaches. First, is the generic approach that could be used as a universal prioritisation criterion for all types of elective surgeries (10, 39, 45). Secondly, the prioritisation criteria were specifically based on measures related to the particular clinical status and recommended for managing the queue for a particular surgery or surgical specialty: orthopaedic surgery (27, 30, 35-37), general surgery (29), neurosurgery (33, 34), bariatric surgery (38, 42), eye surgery (32) and plastic surgery (31).

The goals of a prioritisation methodology should be achieved with scientifically valid prioritisation tools with a transparent mechanism (46). The presented studies have variable risks of bias at varying stages, due to deficiencies in their methods (Table 2) and their results are mixed. The total joint replacement surgery patients that were prioritised by the MAPT (Multi attribute Prioritisation Tool) had improved clinical outcomes with a significantly shorter waiting time (30). Similarly, prioritising patients for cataract surgery using the NIKE (National Indications model for Cataract Extraction) tool reduced waiting times for surgery for those with the greatest need (40). One included study reported on variations of prioritisation among different medical disciplines, which was interpreted mostly as the result of the differences in clinical judgment in various clinical scenarios (33). None of the studies investigated vertical equity approaches of prioritising methods to re-order the waiting list for different elective surgeries using different weights across disciplines, besides vertical equity is often justified by clinical urgency (47). However, standardised universal prioritisation tools were recommended in one study to maintain vertical equity, which would re-order the queue for different specialties in the theatre time allocated to different elective surgery lists (10).

The adverse health outcomes due to long waiting lists for elective surgery are a general policy issue in many countries (48-50). Many of the prioritisation-related interventions that were tested in included studies, were aimed to implement in regional levels or as health system-wide approaches (10, 27, 31, 33, 36-40, 42) while a smaller number of studies focused on single surgery units or individual hospitals (29, 30, 32, 34, 35).

Prioritisation tools based on objective measures of disease severity scales are known as Clinical Priority Assessment Criteria (CPAC) and these have been commonly used in public hospitals (51). Irrespective of whether an explicit CPAC based on clinical parameters was available, it was reported that the order of patients in the surgery list was nearly the same, since clinicians have naturally used their own best judgment to order the queue for the surgery, based on clinical urgency (37). This reflects that clinical parameters alone will not demonstrate the pure urgency and suffering of the respective patient’s condition.

Previous research has identified many potential structural barriers to equitable access to elective surgical care (51) and the importance of prioritisation to the fair allocation for services. Longer waiting times have shown higher detrimental effects to people in lower socio-economic categories (52). Consideration of such equity principles, moral considerations and socio-economic parameters of the patients were suggested as ways to determine the genuine priority access for elective surgery in some included studies (10, 35). The importance of considering moral concerns of patients for prioritisation has also been highlighted for non–life-threatening conditions, such as elective plastic surgeries (31). The added advantage of considering the patients’ perspectives minimises discrimination and leads to patient-centred operating systems.

Some studies included in this review showed that multi-attributes in the prioritisation tools tested for the selected surgeries had a strong association with shorter waiting times along with better post-surgical treatment outcomes (17, 30).

This review also explored different practical approaches to obtain objective measurements for priority scoring. To avoid the limitations of subjective assessments, some studies have recommended the combination of clinician judgment with investigations, reports and details of patient-reported concerns to develop a fair prioritisation tool (10, 31). Instead of the surgeon’s assessment of the clinical or anatomical features of the patient, patient-reported perceived health concerns (40) might indicate a greater need for surgery to a particular patient than others in the queue.

Manipulations and resistance for implementing priority-scoring systems by clinicians have been reported in some studies (51, 53). Similarly, one included study reported that the doctors had not complied with a simple clinical priority guideline which was implemented at the procedural level (39). This indicates the need to convince clinicians on the relative importance of prioritisation and the need to include non-medical factors when determining access to rationed services.

One of the systematic reviews that we included assessed the factors of prioritisation of elective surgical patients in a single surgeon’s waiting list, where the same surgery is managed by many surgeons with multiple surgical lists (41). Unacceptable variability among waiting times in the same specialty in different surgeons or centres and variability in waiting times among different specialties need to be balanced to achieve horizontal and vertical equity for access to health care (44). Adapting prioritisation principles at earlier stages, when the patients were referred to surgical clinics has also been shown to be effective in providing timely services (54). This information might help healthcare managers and policy makers to enhance the local applicability of the implemented prioritisation tools and methods.

## LIMITATIONS OF THIS REVIEW

Our review focused solely on reducing waiting times for elective surgeries and did not include results for patient prioritisation methods used for other purposes, which might also inform decision about strategies for waiting times for elective surgeries. Although there may be multiple purposes, most literature suggests that reducing waiting times for patients in most need is the primary purpose of using prioritisation principals to reorder the queue. Limiting the eligible articles to those published in the six years from 2014-2019 means that we may have failed to include earlier studies that would provide useful evidence. However, relying on recent studies increases the applicability of the results to contemporary practice in healthcare systems. The review includes observational studies, which limits the strength of our recommendations about the effects of interventions, but in the absence of randomised trials, these provide the best available evidence. The included studies assessed prioritisation measures focused on a variety of elective surgery specialties and we did not attempt to summarise the given prioritisation parameters to individual surgeries. Instead, we aimed to provide a holistic approach for prioritisation policies, which could be customised to adapt to any healthcare system. Finally, we have not been able to use the quantitative results of the studies to present meta-analyses of the effects of the interventions or to determine if publication bias has impacted on our conclusions, because of the heterogeneity among the studies.

## CONCLUSIONS

Considering all the quality appraised evidence in the 17 included studies, we have found that this suggests that strategies that support the prioritisation methods suggest can achieve faster access for the patients on waiting lists who are in most need of the surgical care. Having standardised specific prioritisation tools for each specialty is more likely to be effective for horizontal equity by re-ordering the queue of patients waiting for a particular surgery list, but is unlikely to impact on vertical equity across different types of surgery or condition. In addition to clinical assessments, incorporating socio-economic parameters and patients’ moral considerations into prioritisation scoring systems is more effective and more likely to avoid system-associated discrimination in certain surgical specialties. It is challenging to formulate a transparent and consistent national prioritisation system for elective surgeries, but an explicit prioritisation tool with a transparent and objective scoring system based on clear evidence-based criteria might reduce the waiting time for elective surgery.

In summary, this review has listed some factors in a framework (Figure 4) that would address the most important questions asked by healthcare managers and policy makers when seeking a fair prioritisation system for elective surgeries to overcome limitations with local variations. This may be especially important given the impact of the COVID-19 pandemic on elective surgery waiting lists in many countries, and the depletion of resources for routine health care.

**Figure 4.**
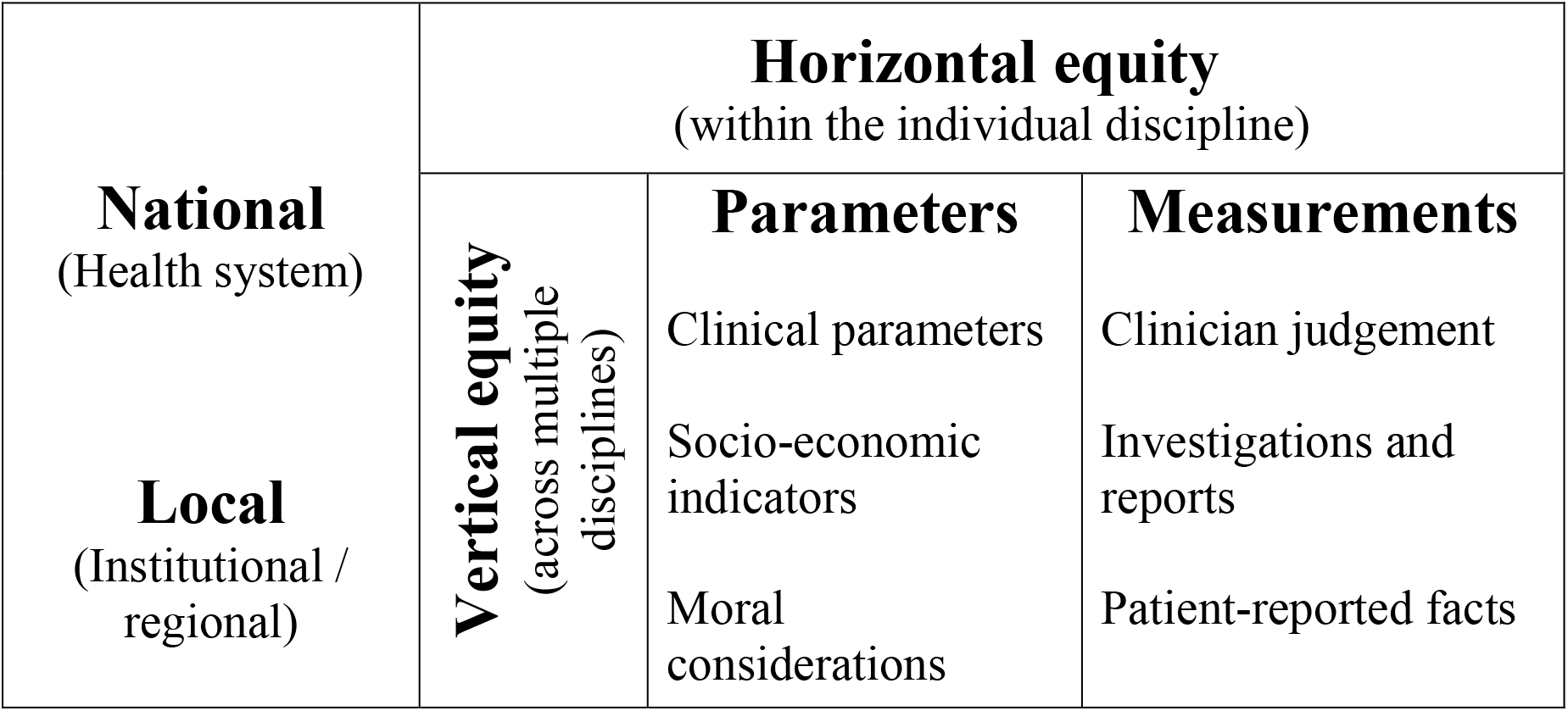
Framework for suggested parameters to be considered in an efficient patient prioritisation system.

However, more research is needed, ideally in the form of randomised trials to quantify the effects of these interventions, as well as economic evaluations leading more precise evidence-informed decision-making.

## Data Availability

The additional data/information will be published as online supplementary files.

<u>Ethics approval and consent to participate</u>: Not applicable, given that this is a systematic review. We registered the review in PROSPERO (CRD42019158455) and reported it in accordance with the PRISMA statement.

<u>Availability of data and materials:</u> Supplementary online file 1, 2, and 3 are attached for online display.

<u>Funding</u>: This project is conducted as part of a self-funded PhD project of the corresponding author.

